# Survival Following Diagnosis of HIV-Associated Kaposi Sarcoma Among Adults in East Africa in the “Treat-All” Era

**DOI:** 10.1101/2024.08.26.24312536

**Authors:** Helen Byakwaga, Aggrey Semeere, Miriam Laker-Oketta, Naftali Busakhala, Esther Freeman, Elyne Rotich, Megan Wenger, Philippa Kadama-Makanga, Job Kisuya, Matthew Semakadde, Bronia Mwine, Charles Kasozi, Bwana Mwebesa, Toby Maurer, David V. Glidden, Kara Wools- Kaloustian, Andrew Kambugu, Jeffrey Martin

## Abstract

**Background:** Despite widespread access to antiretroviral therapy (ART) in the “Treat All” era, HIV-associated Kaposi sarcoma (KS) remains among the most common malignancies in sub-Saharan Africa. Survival after KS diagnosis has historically been poor in Africa, but knowledge whether survival has changed at the population level in the contemporary era has been limited by lack of community-representative surveillance and monitoring systems.

**Methods:** We identified all adult persons living with HIV (PLWH) with a new diagnosis of KS made between 2016 and 2019 during outpatient or inpatient care at prototypical primary care-providing medical facilities in Kenya and Uganda using rapid case ascertainment. Participants were subsequently followed for vital status, including community tracking for those who became lost to follow-up.

**Findings:** Among 411 participants with newly diagnosed KS, 71% were men, median age was 34 (IQR: 30 to 41) years, and 91% had ACTG T1 tumor extent. Over a median follow-up of 7.8 (IQR: 2.4 to 17.9) months, cumulative incidence of death (95% CI) at months 6, 12 and 18 were 34% (30% to 39%), 41% (36% to 46%) and 45% (40% to 51%), respectively. Having the highest number of anatomic sites (11 to 16) harboring KS lesions (hazard ratio 2.2 (95% CI: 1.3-3.8) compared to 1 to 3 sites) and presence of oral KS lesions (hazard ratio 2.2 (95% CI: 1.4-3.3)) were independently associated with higher mortality. Lower hemoglobin and CD4 count as well as higher plasma HIV RNA were also associated with higher mortality.

**Interpretation:** Among PLWH with newly diagnosed KS in East Africa in the “Treat All” era, survival was poor and related to mucocutaneous extent of KS. The findings emphasize the need for better control of KS in Africa, including novel approaches for earlier detection, better linkage to oncologic care, and more potent therapy.

## Introduction

In sub-Saharan Africa, the intersection between the epidemic of HIV infection and what was later known to be endemic transmission of Kaposi sarcoma-associated herpesvirus (KSHV) resulted in an explosion in the incidence of HIV-associated Kaposi sarcoma (KS).^1,2^ After several decades of high KS incidence, widespread use of antiretroviral therapy (ART) has finally begun to curb the frequency of new KS diagnoses in the region,^3^ although impoverished health care systems (with limited capacity to diagnose KS)^4^ and absence of surveillance systems (for enumerating KS)^5^ limit our understanding of the true magnitude of change. Even with widespread ART availability and probable under-reporting of cases, KS remains one of the most frequently reported cancers in the general population in sub-Saharan Africa. It is the second most commonly reported cancer in men and third most common in women.^6^

In addition to being common in sub-Saharan Africa, KS has — prior to the availability of ART — also been deadly.^7,8^ Survival after diagnosis is a critical parameter in cancer epidemiology that captures both the biologic aggressiveness of a particular cancer and the effectiveness of the healthcare system in the management of patients with the cancer. Our understanding of survival following a diagnosis of KS in sub-Saharan Africa in the current “Treat All” ART era^9^ has largely been shaped by extrapolation from resource-rich settings,^10^ clinical trials that have been performed with narrowly-defined populations not representative of real-world populations,^11,12^ or from patients in tertiary care settings.^13–15^ The latter are unlikely to be representative of all cases of KS arising from the community both in terms of severity of disease and socioeconomic status, both of which influence survival. Where survival data outside of clinical trials are available, they often have considerable loss to follow-up.^13–18^ These limitations have collectively severely hampered our ability to understand contemporary survival after diagnosis of KS in the target population in which it is most relevant — all KS that arises from the community.

To address deficiencies in our knowledge regarding contemporary KS survival, we identified, using rapid case ascertainment (RCA), all newly diagnosed cases of HIV-associated KS that arose at several prototypic community-based primary care facilities in Kenya and Uganda.^19^ Participants were subsequently assiduously followed for vital status with scant research-level intervention. We sought to describe real-world survival and determinants of survival after diagnosis of KS among a community-representative sample of persons living with HIV (PLWH) in East Africa in the current “Treat All” era.

## Methods

### Overall Design

We conducted a prospective cohort study of PLWH newly diagnosed with KS. We used a variety of approaches^19^ to scour several community-based health facilities in Kenya and Uganda for all new diagnoses of KS made therein among PLWH. After initial rapid characterization, these patients were then followed to comprehensively monitor vital status, and ultimately, discover determinants of survival (Figure 1).

**Figure 1.**
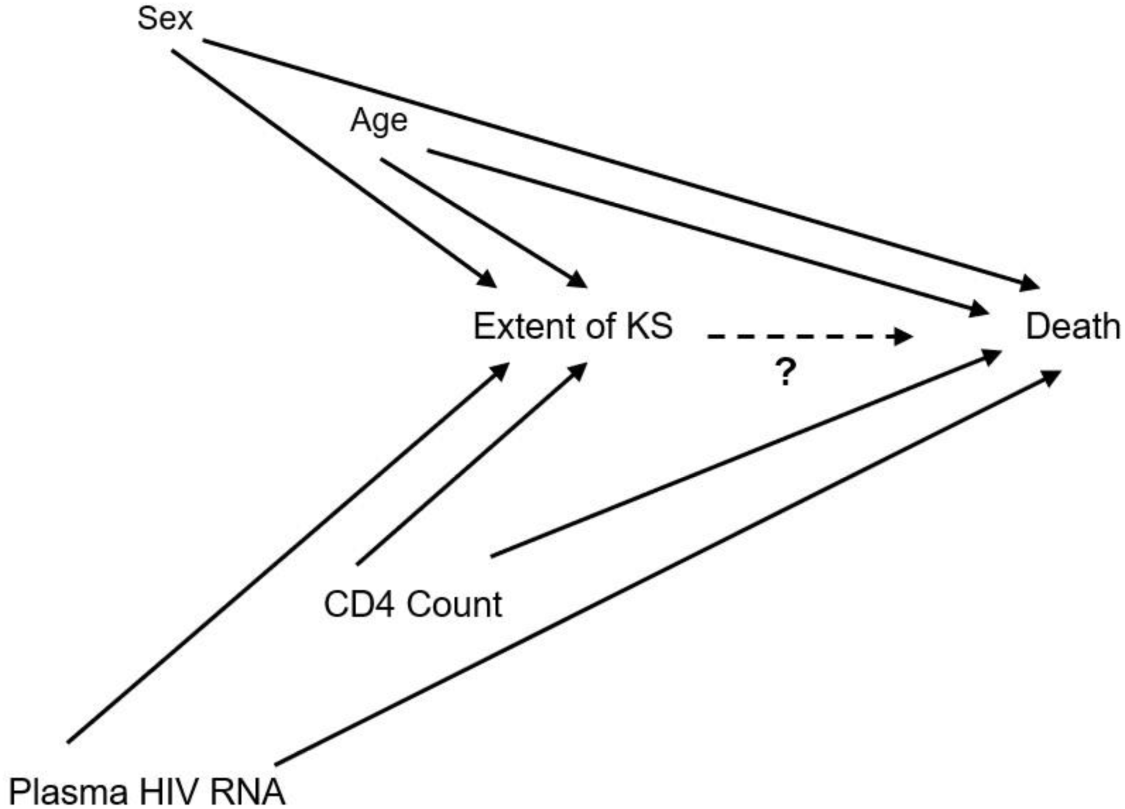
A directed acyclic graph (DAG)^23,24^ depicting our hypothesized conception of the system under investigation. We seek to estimate the causal contribution of age, biologic sex, peripheral CD4+ T cell count, HIV burden (proxy measured by levels of HIV RNA in plasma), and extent of Kaposi sarcoma (KS) on the occurrence of death among adults living with HIV in Kenya and Uganda after the diagnosis of KS. The DAG was used to guide which variables to control for when assessing the total or direct effect of the various constructs on death.In this rendition of the DAG, as depicted by the hashed edge and “?”, we are focusing on investigating the relationship between extent of KS and death. Other perspectives on this DAG were used to investigate potential causal roles for the other variables (e.g., CD4+ T cell count).

### Study Population

*Study sites.* We included consecutive adult (age ≥18 years) PLWH with a new diagnosis of KS made at participating sites: a) The Academic Model Providing Access to Healthcare (AMPATH) network in western Kenya, which was caring for 70,450 PLWH when we began in 2016; b) Masaka Regional Referral Hospital (Masaka-RRH), in Masaka, Uganda, which was caring for 14,380 PLWH when we began in 2018; and c) Mbarara Regional Referral Hospital (Mbarara-RRH) clinic in Mbarara, Uganda, which was caring for 10,800 PLWH when we began in 2018. These sites were chosen because they offered what we believed was the best opportunity to investigate a community-representative sample of new diagnoses of HIV-associated KS in Kenya and Uganda. In addition to providing primary care to large populations of PLWH (documented with searchable electronic medical record-derived databases), each site offered free-of-charge skin biopsies for patients with suspected KS.^19^ These biopsy services attract referrals not only from existing patients at the outpatient and inpatient units within the participating facilities, but also from other persons originating from the community who may have been out of care or receiving care at facilities without biopsy capability.

*Identification of study participants.* Effort was made to identify and evaluate all KS diagnoses made in PLWH as soon as they occurred at each site using the RCA approach.^19^ RCA refers to the expeditious and thorough evaluation of a patient shortly following diagnosis of a particular disease; it is most commonly used in rapidly progressive or fatal diseases.^20,21^ In order to identify KS diagnoses as soon as they occurred at the participating facilities, we performed weekly searches of the skin biopsy service records, electronic medical records of the HIV primary care clinics, adult inpatient medical wards, histopathology laboratory records, and, depending on the site, dermatology and oncology clinic records. In addition, we obtained ad hoc phone and in-person notifications from clinicians regarding new suspected KS cases. KS diagnosis was confirmed on histopathology performed at the respective local pathology laboratories. If lesions were deemed unsafe to biopsy (e.g., conjunctival or oral locations), a clinical diagnosis of KS was made based on the characteristic macroscopic appearance. After identification of eligible patients, effort was then made to rapidly contact them to make detailed measurements at what was known as the RCA encounter.

### Measurements

*Questionnaire-based*. At the RCA encounter, we used interviewer-administered questionnaires to document socio-demographic characteristics and medical history. Following enrollment, participants were evaluated every four months with in-person or phone-administered interviews to vital status. We intentionally limited measurements during follow-up to minimize research-level intervention. Community visits were performed using methods we previously established^22^ to trace participants who did not return for scheduled study visits and could not be reached by phone.

*Physical examination.* Cutaneous and oral examinations were performed. The extent of KS was evaluated by ascertaining number of anatomic sites with cutaneous lesions suspected to be KS, the total number of KS-suspicious lesions, presence of edema not otherwise explained, and presence of KS-suspicious lesions that had macroscopic tumor morphology.

*Laboratory*. Complete blood count, plasma HIV RNA level (Amplicor HIV Monitor version 1.5 or the Cobas Taqman HIV-1 version 1.0 assays; Roche, Branchburg, NJ) and CD4+ T-cell counts (FACSCalibur; Becton Dickinson, San Jose, CA) were measured at the respective clinical laboratories of the participating sites.

### Statistical Analysis

To estimate survival after KS diagnosis, we used the Kaplan-Meier approach and included all adult PLWH with a new diagnosis of KS identified at the participating sites during the study period. Time zero for a given patient was the date when the health system was first certain about the KS diagnosis. This was the date of histopathological diagnosis (for those with biopsy) or the date when the health system made a clinical KS diagnosis for those in whom a biopsy could not performed because of safety concerns. In the base-case analysis, we defined the observation time as duration from time zero to the date of death for those known to have died with known death dates. For those known to have died but without precise knowledge of death dates, we used the median duration to death obtained for those with known dates of death. For individuals not known to have died, we defined observation time as duration from time zero to last date known to be alive. We also performed two sensitivity analyses. First, we assumed all those lost to follow-up had died the day after their last known date alive. Second, we assumed all those lost to follow-up were still alive as of October 01, 2019, the date of administrative censoring.

To assess determinants of death, we only included participants who had detailed measurements performed, through the RCA mechanism, within 30 days of KS diagnosis. This analytic sample thus represented a subset of the entire study sample that was used for the descriptive survival estimate. We used proportional hazards regression to evaluate determinants of death. Directed acyclic graphs (DAGs) were used to inform which variables to adjust for to manage confounding (Figure 1).^23,24^ For missing data, we performed multiple imputation with iterative chained equations^25^ and included the Nelson–Aalen estimate of the cumulative hazard of death.^26^ Scaled Schoenfeld residuals were used to examine the proportional hazards assumption.^27^ All analyses were performed using Stata 15 (College Station, TX).

## Results

### Characteristics of Adults with Newly Diagnosed HIV-Associated KS

Between July 1, 2016 and July 31, 2019, we identified 411 adult PLWH with newly diagnosed KS. Of these, 269 (65%) were identified at AMPATH, 85 (21%) at Masaka-RRH, and 57 (14%) at Mbarara-RRH. The majority (71%) were men, and the median (interquartile range, IQR) age was 34 (30-41) years. Characteristics for the 411 were similar to the 262 participants for whom we performed detailed clinical assessments within 30 days of a new KS diagnosis (Figure 2). Among these 262 participants, 171 (65%) were identified at AMPATH, 66 (25%) at Masaka-RRH, and 25 (10%) at Mbarara-RRH. At KS diagnosis, the majority (69%) of these 262 participants were male, the median (IQR) age was 35 (30-41) years, hemoglobin was 10.6 (8.8 to 12.7) g/dl and CD4 count was 233 (84-403) cells/µl (Table 1). Almost all (98%) participants reported using ART; 40% had undetectable plasma HIV RNA (<40 copies/ml), and 22% had >10,000 copies/ml. KS diagnosis was based on histopathology except for five participants who only had KS-suspicious oral lesions that were deemed unsafe to biopsy. The median number of anatomic sites with KS lesions was six (IQR: 4 to 10), 77% had edema, 35% had oral KS lesions, 35% had at least one KS-suspicious lesion with macroscopic tumor morphology, and most (91%) had advanced KS by virtue of T1 AIDS Clinical Trials Group (ACTG) stage.

**Figure 2.**
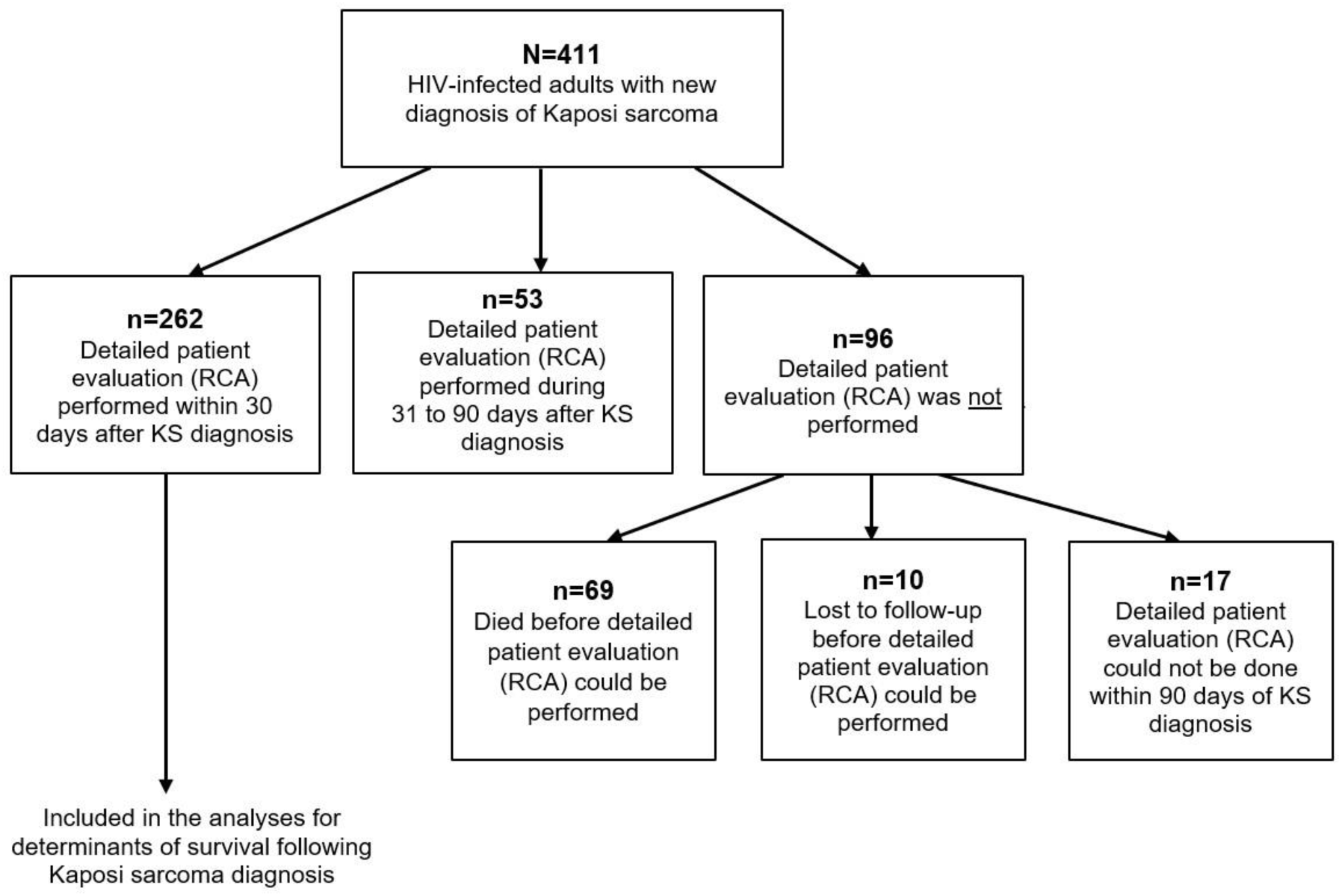
Schema of study of adults living with HIV infection and newly diagnosed with Kaposi sarcoma (KS) at one of three participating sites in Kenya and Uganda who were included in this study. The entire study population was used to derived descriptive estimates of mortality of after diagnosis of KS. RCA refers to rapid case ascertainment, which is the process of attempting to perform rapid identification and thorough research-level evaluation of a patient shortly following the diagnosis of a disease. Only participants who had RCA performed within 30 days of KS diagnosis were included in analytic study of determinants of mortality.

**Table 1.** Sociodemographic and clinical characteristics at the time of diagnosis of Kaposi sarcoma (KS) among adults living with HIV who were newly diagnosed with KS in Kenya and Uganda from 2016 to 2019.

| Characteristic | Value* (N=262) |
| --- | --- |
| **Age, years** | 35 (30 to 41) |
| **Male sex** | 69% |
| **Highest level of education completed** |  |
| < Primary education | 39% |
| Primary completed, but secondary education not completed | 40% |
| Completed secondary, vocational or tertiary education | 21% |
| **Monthly income, U.S. dollars** | 79 (31-168) |
| **Number of distinct KS-suspicious skin or oral lesions** |  |
| 1-49 | 63% |
| > 50 | 37% |
| **Number of anatomic sites<sup>†</sup> with KS-suspicious lesions** | 6 (4 to 10) |
| **Presence of KS-suspicious oral lesions** | 35% |
| **Presence of KS-suspicious lesions with tumor morphology** | 35% |
| **Presence of edema attributed to KS** | 77% |
| **ACTG Stage T1<sup>‡</sup>** | 91% |
| **History of tuberculosis or cryptococcal meningitis** | 23% |
| **Hemoglobin, g/dl** | 10.6 (8.8 to 12.7) |
| **CD4+ T cells, count/μl** |  |
| ≤ 50 | 17% |
| 51-200 | 26% |
| 201-500 | 41% |
| > 500 | 16% |
| **HIV RNA, copies/ml** |  |
| ≤ 40 | 40% |
| 41-1,000 | 27% |
| 1001-10,000 | 11% |
| > 10,000 | 22% |

\* median (interquartile range) unless indicated
<sup>†</sup> Anatomic sites for KS include: head, oral cavity, neck, chest, abdomen, back, right arm, left arm, right leg, left leg, right hand, left hand, right foot, left foot, genital and gluteal (total = 16 anatomic sites)
<sup>‡</sup> ACTG T1 stage was defined as having any of the following based on physical exam or self-reported history: tumor-associated edema or ulceration; extensive oral KS lesions; visceral KS<sup>32</sup>

### Mortality among Adults with Newly Diagnosed HIV-Associated KS

Over a median follow-up of 7.8 (IQR: 2.4 to 17.9) months, a total of 174 deaths occurred among the 411 participants with a new diagnosis of KS. For five participants for whom the date of death could not be confirmed, we assumed observation time to be 58 days (the median time to death for all deaths with known dates of death). The cumulative incidence of death (95% CI) at months 6, 12 and 18 following KS diagnosis was 34% (30% to 39%), 41% (36% to 46%) and 45% (40% to 51%), respectively (Figure 3). Vital status of only 10 (2.4%) patients could not be ascertained because the study team was unable to reach them by phone or track them in the community. With so few patients lost to follow-up, sensitivity analyses were unsurprisingly consistent with the base-case analysis. Considering the worst-case scenario, cumulative incidence of death (95% CI) at months 6, 12 and 18 was 36% (32% to 41%), 43% (38% to 48%) and 47% (42% to 53%), respectively. For the best-case scenario, it was 33% (29% to 38%), 40% (35% to 45%) and 44% (39% to 49%), respectively.

**Figure 3.**
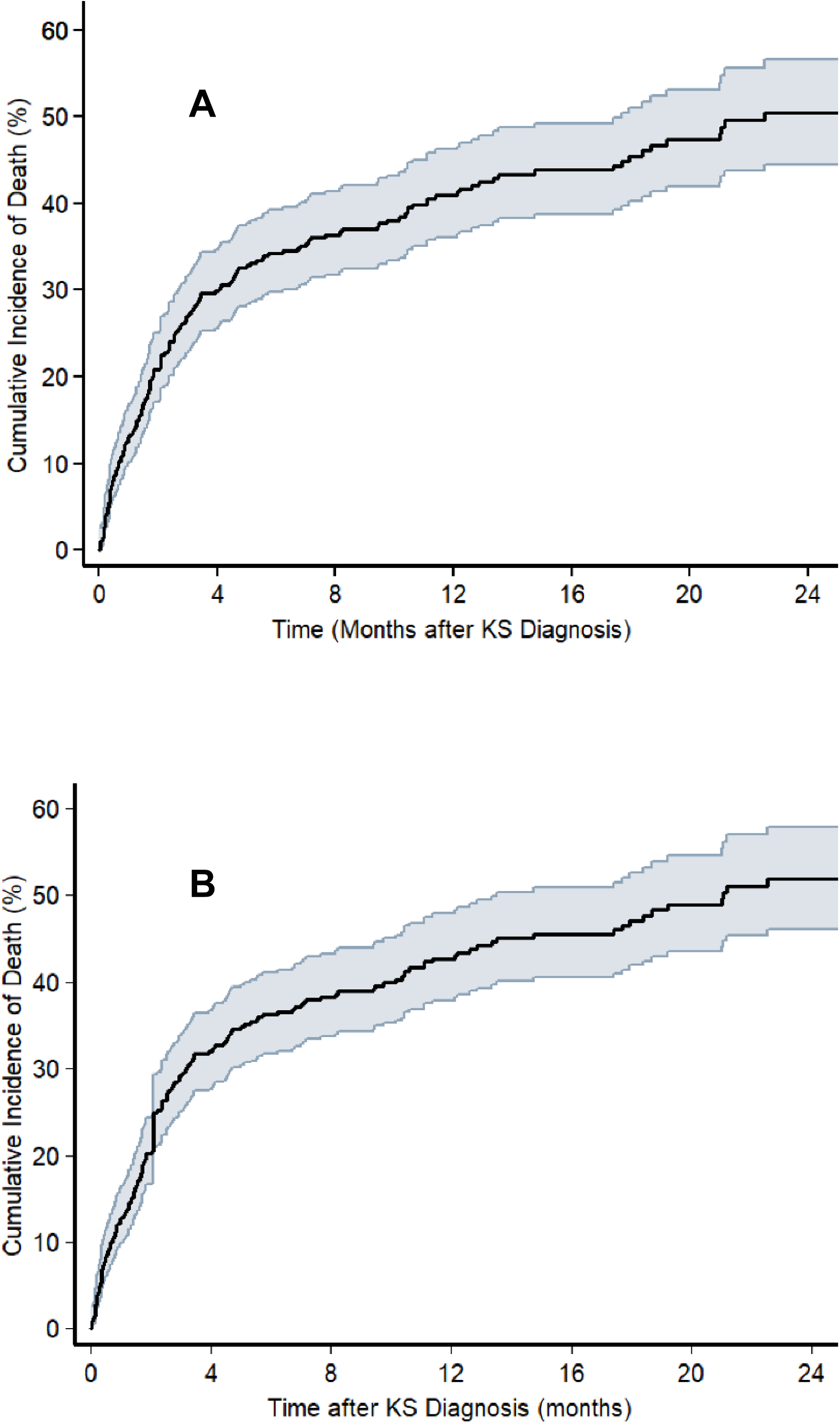
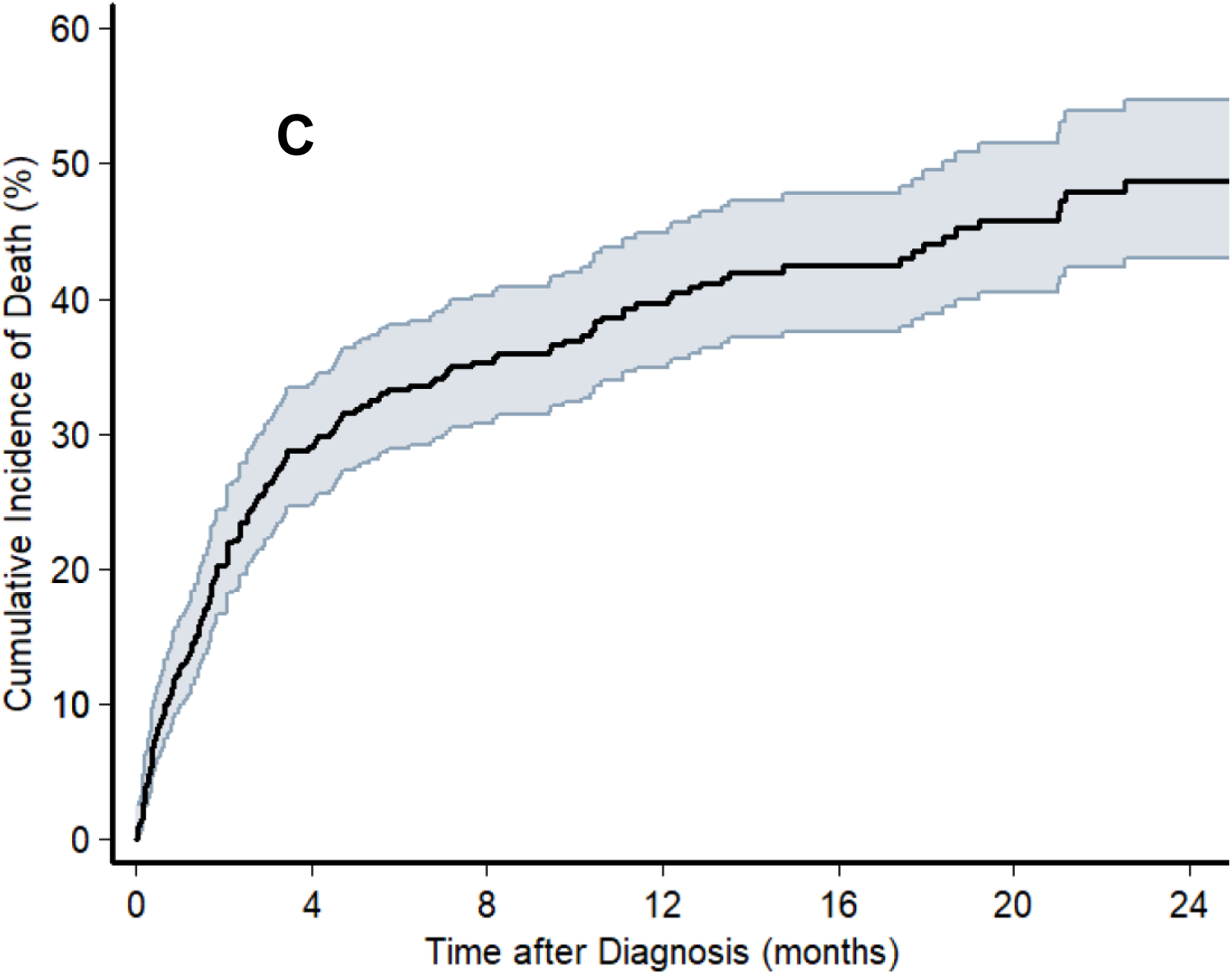
Cumulative incidence of death, as determined by the Kaplan-Meier estimator, following diagnosis of KS among adults living with HIV with newly diagnosed Kaposi sarcoma between 2016 and 2019 at one of three health systems in Kenya and Uganda. The shaded area represents the 95% confidence interval around the cumulative incidence estimate. Panel A depicts the base-case analysis in which participants who became lost to follow-up were censored on the last date they were known to be alive. Panel B depicts a sensitivity analysis in which participants who became lost to follow-up were classified as dead the day after they were last known to be alive (“worst case” mortality). Panel C depicts a sensitivity analysis in which participants who became lost to follow-up were classified as alive as of the date of administrative censoring for the study population (“best case” survival).

### Determinants of Mortality among Adults with HIV-Associated KS

There was no strong evidence of an association between socioeconomic or demographic characteristics (age, sex, education, or income) at the time of new KS diagnosis and subsequent mortality (Table 2). Regarding medical history, individuals with a previous history of opportunistic infection (tuberculosis or cryptococcal meningitis) had a higher mortality compared to those who did not in unadjusted analyses, but this was no longer statistically significant after adjustment for sex, age, CD4 count, and plasma HIV RNA level.

**Table 2.**
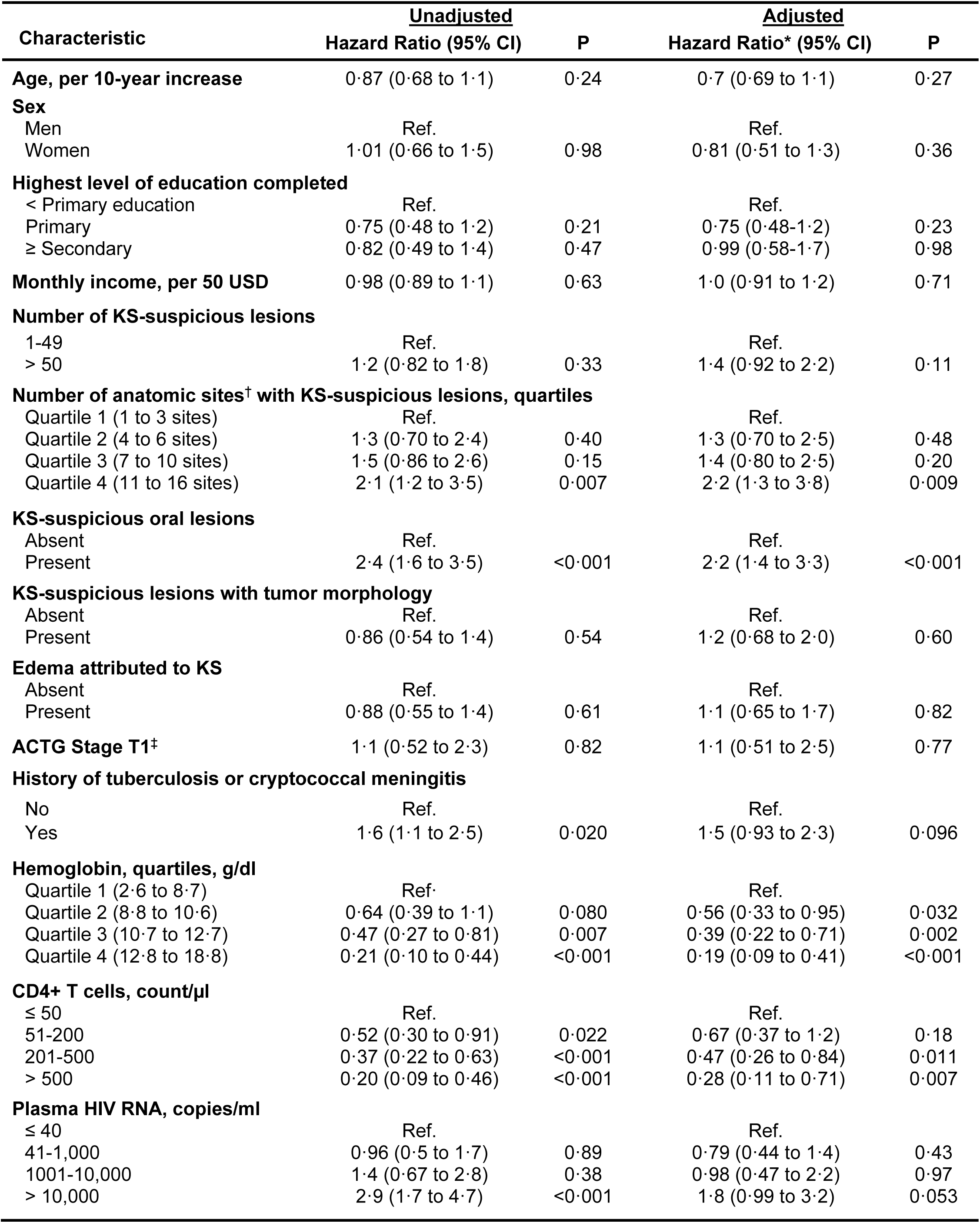
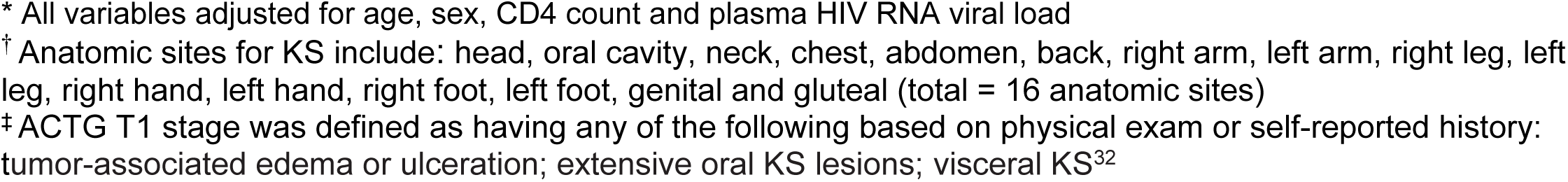
Estimation of the association between a variety of characteristics at the time of diagnosis of Kaposi sarcoma (KS) and subsequent time-to-death among adults living with HIV who were newly diagnosed with KS in Kenya and Uganda between 2016 and 2019.

| Characteristic | <u>Unadjusted</u> |  | <u>Adjusted</u> |  |
| --- | --- | --- | --- | --- |
|  | Hazard Ratio (95% CI) | P | Hazard Ratio* (95% CI) | P |
| **Age, per 10-year increase** | 0.87 (0.68 to 1.1) | 0.24 | 0.7 (0.69 to 1.1) | 0.27 |
| **Sex** |  |  |  |  |
| Men | Ref. |  | Ref. |  |
| Women | 1.01 (0.66 to 1.5) | 0.98 | 0.81 (0.51 to 1.3) | 0.36 |
| **Highest level of education completed** |  |  |  |  |
| < Primary education | Ref. |  | Ref. |  |
| Primary | 0.75 (0.48 to 1.2) | 0.21 | 0.75 (0.48-1.2) | 0.23 |
| ≥ Secondary | 0.82 (0.49 to 1.4) | 0.47 | 0.99 (0.58-1.7) | 0.98 |
| **Monthly income, per 50 USD** | 0.98 (0.89 to 1.1) | 0.63 | 1.0 (0.91 to 1.2) | 0.71 |
| **Number of KS-suspicious lesions** |  |  |  |  |
| 1-49 | Ref. |  | Ref. |  |
| > 50 | 1.2 (0.82 to 1.8) | 0.33 | 1.4 (0.92 to 2.2) | 0.11 |
| **Number of anatomic sites<sup>†</sup> with KS-suspicious lesions, quartiles** |  |  |  |  |
| Quartile 1 (1 to 3 sites) | Ref. |  | Ref. |  |
| Quartile 2 (4 to 6 sites) | 1.3 (0.70 to 2.4) | 0.40 | 1.3 (0.70 to 2.5) | 0.48 |
| Quartile 3 (7 to 10 sites) | 1.5 (0.86 to 2.6) | 0.15 | 1.4 (0.80 to 2.5) | 0.20 |
| Quartile 4 (11 to 16 sites) | 2.1 (1.2 to 3.5) | 0.007 | 2.2 (1.3 to 3.8) | 0.009 |
| **KS-suspicious oral lesions** |  |  |  |  |
| Absent | Ref. |  | Ref. |  |
| Present | 2.4 (1.6 to 3.5) | <0.001 | 2.2 (1.4 to 3.3) | <0.001 |
| **KS-suspicious lesions with tumor morphology** |  |  |  |  |
| Absent | Ref. |  | Ref. |  |
| Present | 0.86 (0.54 to 1.4) | 0.54 | 1.2 (0.68 to 2.0) | 0.60 |
| **Edema attributed to KS** |  |  |  |  |
| Absent | Ref. |  | Ref. |  |
| Present | 0.88 (0.55 to 1.4) | 0.61 | 1.1 (0.65 to 1.7) | 0.82 |
| **ACTG Stage T1<sup>‡</sup>** | 1.1 (0.52 to 2.3) | 0.82 | 1.1 (0.51 to 2.5) | 0.77 |
| **History of tuberculosis or cryptococcal meningitis** |  |  |  |  |
| No | Ref. |  | Ref. |  |
| Yes | 1.6 (1.1 to 2.5) | 0.020 | 1.5 (0.93 to 2.3) | 0.096 |
| **Hemoglobin, quartiles, g/dl** |  |  |  |  |
| Quartile 1 (2.6 to 8.7) | Ref. |  | Ref. |  |
| Quartile 2 (8.8 to 10.6) | 0.64 (0.39 to 1.1) | 0.080 | 0.56 (0.33 to 0.95) | 0.032 |
| Quartile 3 (10.7 to 12.7) | 0.47 (0.27 to 0.81) | 0.007 | 0.39 (0.22 to 0.71) | 0.002 |
| Quartile 4 (12.8 to 18.8) | 0.21 (0.10 to 0.44) | <0.001 | 0.19 (0.09 to 0.41) | <0.001 |
| **CD4+ T cells, count/μl** |  |  |  |  |
| ≤ 50 | Ref. |  | Ref. |  |
| 51-200 | 0.52 (0.30 to 0.91) | 0.022 | 0.67 (0.37 to 1.2) | 0.18 |
| 201-500 | 0.37 (0.22 to 0.63) | <0.001 | 0.47 (0.26 to 0.84) | 0.011 |
| > 500 | 0.20 (0.09 to 0.46) | <0.001 | 0.28 (0.11 to 0.71) | 0.007 |
| **Plasma HIV RNA, copies/ml** |  |  |  |  |
| ≤ 40 | Ref. |  | Ref. |  |
| 41-1,000 | 0.96 (0.5 to 1.7) | 0.89 | 0.79 (0.44 to 1.4) | 0.43 |
| 1001-10,000 | 1.4 (0.67 to 2.8) | 0.38 | 0.98 (0.47 to 2.2) | 0.97 |
| > 10,000 | 2.9 (1.7 to 4.7) | <0.001 | 1.8 (0.99 to 3.2) | 0.053 |

\* All variables adjusted for age, sex, CD4 count and plasma HIV RNA viral load
† Anatomic sites for KS include: head, oral cavity, neck, chest, abdomen, back, right arm, left arm, right leg, left leg, right hand, left hand, right foot, left foot, genital and gluteal (total = 16 anatomic sites)
‡ ACTG T1 stage was defined as having any of the following based on physical exam or self-reported history: tumor-associated edema or ulceration; extensive oral KS lesions; visceral KS<sup>32</sup>

A higher number of anatomic sites with mucocutaneous KS-suspicious lesions was associated with higher mortality in both unadjusted and adjusted analyses. Mortality in individuals with 11 to 16 sites harboring KS-suspicious lesions (the highest quartile) was 2.2 (95% CI: 1.3 to 3.8) times as high as in those with 1 to 3 sites involved (the lowest quartile) even after adjustment for age, sex, CD4 count, and plasma HIV RNA level. In addition, presence of oral KS lesions was associated with higher mortality in both unadjusted and adjusted analyses. There was no strong evidence for an association between total number of cutaneous KS lesions (dichotomized at 50 lesions), presence of edema, presence of KS tumor lesions or having ACTG stage T1 KS disease and death.

Higher CD4 counts were protective against death in a dose-response manner. The hazard ratio comparing participants with CD4 counts >500 and 201-500 cells/µl to those with CD4 counts ≤50 cells/µl was 0.28 (95% CI: 0.11 to 0.71) and 0.47 (95% CI: 0.26 to 0.84), respectively, even after adjustment for age, sex, and plasma HIV RNA level. We also observed that a higher plasma HIV RNA level was associated with higher mortality. Participants with plasma HIV RNA levels >10,000 copies/ml had 1.8 times (95% CI: 0.99-3.2) higher rate of mortality than those with ≤40 copies/ml, again after adjustment for age, sex and CD4 count. Finally, there was also a monotonic relationship between hemoglobin and mortality. In analyses adjusted for sex, age, CD4 count, and plasma HIV RNA level, individuals with the highest levels of hemoglobin (quartile 4) had 81% (95% CI: 59% to 91%) lower rate of mortality than those with the lowest levels of hemoglobin (quartile 1).

## Discussion

In late 2015, the WHO recommended initiation of ART in all PLWH, regardless of CD4 count or clinical stage, the dawning of the “Treat All” era.^9^ At the most macroscopic level, “Treat All” has been successful in most resource-limited settings, diminishing all-cause death related to HIV.^28,29^ What is less understood is whether all individual severe HIV-related complications have fared equally in diminution of mortality. Overcoming many of the limitations of prior research in terms of population representativeness and completeness of follow-up, we observed remarkably high mortality in the first six months (and continued mortality through 18 months) following diagnosis of KS in a large sample of PLWH in East Africa in the “Treat All” era. Mucocutaneous extent of KS, as indicated by a higher number of anatomical sites with KS lesions and the presence of oral lesions, as well as conventional biological markers of HIV infection and hemoglobin level were associated with poor survival. These findings establish a benchmark for KS survival (and methodology for how to study it) among PLWH in the current era in sub-Saharan Africa to which future estimates can be compared.

Prior to the availability of effective ART, one-year mortality after a diagnosis of KS in sub-Saharan Africa was approximately 60% to 70%.^7,8^ Previous studies in sub-Saharan Africa in the ART era have suggested a substantial decline in mortality after KS diagnosis, with one-year mortality estimated to be 21% to 28%. ^12–14,17^ There are several reasons that might explain why mortality in our study (41% at one year) was substantially higher than these prior studies. For the previous studies that were clinical trials, patients with severe KS or other abnormalities (e.g., low hemoglobin or elevated liver enzymes) were excluded,^11,17^ and, consequently, individuals with highest risk of death might have been omitted. In addition, patients enrolled in clinical trials are typically more closely monitored than patients in routine care with more treatment availability in the event of concurrent conditions or treatment complications. Among the observational studies, some reports were of patients receiving oncology care at tertiary centers.^14,15,18^ These patients are typically enriched with those with the economic resources to access tertiary care, a factor that limits the representativeness of these patients in comparison to all cases of KS arising from the community. Other reports were based on review of data collected during routine HIV care,^16,17^ a setting in which access to histological confirmation of KS is typically not readily available (unlike in our study), thus calling into question the representativeness of patients who were able to receive a biopsy-confirmed diagnosis of KS. Finally, there was considerable lost to follow-up in many of the observational studies, up to 67%.^13–18^ Losses to follow up may result in underreporting of death and underestimation of mortality.^16,22^ In fact, in prior work based on data collected during routine care, mortality in patients with KS was shown to double when attempts were made to account for patients lost to follow-up.^22^ Given the threat to validity in survival estimates posed by those lost to follow-up, we expended considerable effort to document vital status of our participants, and only 10 were ultimately lost to follow-up.

Extent of disease at diagnosis is an important determinant of survival for many cancers.^30,31^ The ACTG staging system for KS, the most commonly used scheme for KS globally, was developed in resource-rich countries prior to availability of ART.^32^ Within this staging system, patients are classified as advanced KS (T1 stage) based on the presence of tumor-associated edema or ulceration; oral lesions; or visceral KS. The number or extent of cutaneous lesions are notably absent as criteria. Given that ART may modify the presentation or course of KS, it is unclear how well the ACTG staging system performs in the current era.^33,34^ In what we believe is the first such published observation, we found that patients with the greatest anatomic extent of cutaneous KS had poorer survival. Of note, we did not find a role for the presence of edema or ACTG T1 stage.

The relationship between hemoglobin and survival has been previously reported in unselected PLWH on ART, ^35,36^ as well as among patients with HIV-associated KS, in sub-Saharan Africa.^37^ While low levels of hemoglobin may be indicative of HIV disease progression,^38^ we found that the association between hemoglobin and survival was independent of plasma HIV RNA viral load and CD4 count, suggesting either an additional HIV-related mechanism or a direct effect of KS. Specifically, low hemoglobin has been associated with markers of KS inflammation and immune activation,^39,40^ which have been shown to persist despite ART-mediated HIV viral suppression^41,42^ and are associated with higher mortality.^43^ Alternatively, lower levels of hemoglobin may be a consequence and thus marker of gastrointestinal KS.^32^

A limitation of our work is that despite extensive effort to identify all newly diagnosed KS among PLWH at the participating facilities, we might have missed some patients, particularly those receiving clinical KS diagnoses (i.e., no biopsy confirmation) that either were only recorded in isolated clinical venues (e.g., a specialty clinic) or inadvertently never documented. We believe this was uncommon given the easy access to biopsy at the participating sites and the many tentacles we had to scour records in outpatient and inpatient venues. Thus, our study sample is credibly representative of all new KS diagnoses made in the respective surrounding communities. What is more likely is that we missed some KS that arose from the community that was never diagnosed. We are anecdotally aware that some people develop KS and either never present to care (die at home) or present to care and die because of overwhelming comorbid conditions, with their KS never formally diagnosed. This differs substantially from resource-rich settings in which most cancer that occurs is eventually diagnosed and recorded. Thus, we concede that our sample cannot be stated to be truly population-representative of all biologic or clinical occurrences of KS. Indeed, virtually all cancer studies in resource-limited settings, like sub-Saharan Africa, suffer from this threat and often without recognition. Cancer is an entity that requires a highly functional healthcare system to measure it. Without such a system, resource-limited settings suffer from a disparity in the data needed for accurate characterization of their cancer epidemiology.

There are several implications of our findings. First and foremost, the poor survival following a diagnosis of HIV-associated KS in the “Treat-All” ART era in East Africa, coupled with its continued substantial incidence, means that KS remains a clinically relevant condition. Continued work is needed on all fronts of primary, secondary, and tertiary prevention of KS. Second, the high prevalence of advanced KS disease at the time of diagnosis and influence of advanced disease on survival suggest that the greatest impact on survival may arise from early detection of KS. This is especially true given the lack of cure for advanced KS and the difficulty in ensuring availability of curative therapy even if it did exist. Third, contemporary staging systems are needed for HIV-related KS in Africa. We have identified two readily measured variables — extent of cutaneous KS and hemoglobin — that merit replication by others as predictors of survival. Finally, although not directly proven in our work, it is likely that better management of HIV infection among PLWH with KS (i.e., greater achievement of undetectable plasma HIV RNA and higher CD4 count) will improve survival.

In summary, among adult PLWH with newly diagnosed KS, we documented poor survival in the “Treat-All” ART era. The findings are a clarion call for better control of HIV-associated KS in Africa. This includes detecting KS in its early stages and improving access to diagnosis and more potent chemotherapy. In the absence of government-funded registries to monitor cancer survival in sub-Saharan Africa, we hope that other researchers in the region will fill the gap and generate similarly accurate mortality estimates to further monitor KS survival, drawing requisite attention to this disease until a time it is no longer clinically relevant.

### Previous presentation

This work was previously presented in part at the 17th International Conference on Malignancies in AIDS and Other Acquired Immunodeficiencies in Bethesda, Maryland, on October 21-22, 2019.

### Funding

Research reported in this publication was supported by the National Institute of Allergy and Infectious Diseases, the National Cancer Institute, and Fogarty International Center, in accordance with the regulatory requirements of the National Institutes of Health under Award Numbers U01AI069911, U54 CA190153, P30 AI027763, U54 CA25457, P30 CA082103 and K43 TW011987. The funders did not have a role in design of the study, analysis, interpretation of data, or in writing the manuscript. The content is solely the responsibility of the authors and does not necessarily represent the official views of the National Institutes of Health.

### Ethics approval and consent to participate

This study received regulatory approval in both Kenya and Uganda. In Kenya, the study was approved by the Institutional Research Ethics Committee at Moi University (Protocol No. 0001827) and in Uganda by the Makerere University College of Health Sciences School of Biomedical Sciences Higher Degrees Research and Ethics Committee (Protocol No. SBS-HD-REC-495).

### Consent for publication

No individually identifiable data – not applicable.

### Availability of data and materials

The datasets used and analyzed during the current study are available from the corresponding author on reasonable request.

### Authorship contribution statement

*Helen Byakwaga:* Conceptualization, Methodology, Formal analysis, Writing-Original Draft, Project administration, Visualisation. *Aggrey Semeere:* Conceptualization, Methodology, Writing-Review and Editing, Project administration. *Miriam Laker-Oketta:* Methodology, Writing-Review and Editing, Project administration. *Naftali Busakhala:* Project administration, Writing-Review and Editing. *Esther Freeman:* Methodology, Writing-Review and Editing, Project administration. *Elyne Rotich:* Project administration, Writing-Review and Editing. *Megan Wenger:* Software, Data Curation, Writing-Review and Editing, Project administration. *Philippa Kadama-Makanga:* Project administration, Writing-Review and Editing. *Job Kisuya:* Project administration, Writing-Review and Editing. *Matthew Ssemakadde:* Project administration, Data Curation. Bronia Mwine: Project administration, Writing-Review and Editing, Data Curation*. Charles Kasozi:* Project administration, Writing-Review and Editing*. Mwebesa Bwana:* Project administration, Resources. *Toby Maurer:* Methodology, Writing-Review and Editing, Project administration. *David V, Glidden:* Methodology, Writing-Review and Editing. *Kara Wools-Kaloustian:* Funding acquisition, Writing-Review and Editing. *Andrew Kambugu:* Funding acquisition, Supervision. *Jeffrey Martin:* Conceptualization, Methodology, Writing-Review and Editing, Funding acquisition, Validation, Supervision.

## Data Availability

All data produced in the present study are available upon reasonable request to the authors

## Acknowledgements

We thank all the research staff who contributed to data collection at the different health facilities that participated in this study. AMPATH: Celestine Lagat, Raphael Kobilo, Kipkoir Koima and Linda Chemtai; Masaka-RRH: Haruna Semuwemba, Stella Nabunya and Fiona Nassonko; and Mbararra-RRH: Bronia Mwine, Martin Mwebesa and Placidia Oinembabazi.

